# Carotid plaque dynamic contrast-enhanced magnetic resonance imaging normalised signal intensity reproducibly differs between plaque and vessel wall

**DOI:** 10.64898/2026.02.20.26346739

**Authors:** Thomas Readford, Gonzalo J Martínez, Sanjay Patel, Peter L. Kench, Marcelo Andía, Martin Ugander, Nicola Giannotti

## Abstract

**Background:** Dynamic contrast-enhanced magnetic resonance imaging (DCE-MRI) enables non-invasive characterization of carotid atherosclerotic plaque.

**Purpose:** To evaluate the performance and reproducibility of a simplified DCE-MRI quantification method for carotid plaque assessment.

**Methods:** T1-weighted black-blood DCE-MRI of the carotid arteries at 3T was performed at baseline and after six months in patients with mild-to-moderate atherosclerotic lesions in a pilot placebo-controlled randomized trial evaluating the effects of low-dose (0.5mg daily) colchicine therapy on carotid plaque volume. DCE-MRI signal intensity was measured in manually drawn regions of interest in the plaque core, remote non-atherosclerotic vessel wall, and skeletal muscle. Peak signal intensities were normalized to skeletal muscle signal in the same slice.

**Results:** In patients (n=28, median [interquartile range] age 72 [64–74] years, 36% female, n=13/15 colchicine/placebo), normalized peak signal intensity was higher in the plaque core than in the remote vessel wall at both baseline (3.5 [2.3–4.1] vs 2.1 [1.7–2.5], p<0.001) and follow-up (3.2 [2.5–4.4] vs 2.0 [1.7–2.5], p<0.001). Measurements did not differ between baseline and follow-up for all patients (0.7±0.7 for plaque core, 0.6±0.4 for remote vessel wall, p>0.80 for both) nor between colchicine intervention and placebo control (p>0.35 for either region).

**Conclusions:** Normalised peak signal intensity on DCE-MRI was consistently higher in the carotid plaque core than in the remote vessel wall, showed excellent reproducibility in both regions over six months, and was not altered by colchicine treatment. This simplified, muscle-normalised approach may facilitate future studies exploring DCE-MRI measures potentially related to plaque vulnerability.

## INTRODUCTION

Carotid atherosclerotic plaque neovascularisation and permeability have been associated with increased plaque inflammation and instability, potentially causing major adverse cerebrovascular events (MACCE). (1) Dynamic contrast-enhanced magnetic resonance imaging (DCE-MRI) of carotid plaque can non-invasively quantify contrast-related parameters such as signal intensity, k-trans, and time-to-peak that may serve as surrogate markers for the evaluation of biological processes such as plaque neovascularisation and endothelial permeability. (2, 3) However, due to the non-linear nature of uptake into the extracellular space for gadolinium-based contrast agents, current quantitative approaches to carotid plaque DCE-MRI typically rely on complex kinetic modelling of gadolinium-based contrast agent behaviour, limiting their translatability beyond the pre-clinical setting. (4, 5) Today, considerable debate persists regarding the most appropriate model (e.g., Tofts, extended Tofts, or Patlak) for accurately reflecting plaque neovascularisation, permeability, and associated risk of major adverse cardiovascular or cerebrovascular events. (5) Therefore, it may be preferable to have simpler and reproducible DCE-MRI-derived markers that could serve as accessible surrogates for contrast enhancement patterns related to plaque vulnerability, potentially facilitating broader clinical and research applications.

DCE-MRI with kinetic modelling has previously been used to evaluate changes in carotid microvascular permeability in response to lipid-lowering therapies (6). However, using DCE-MRI to assess accessible imaging biomarkers is of interest for identifying contrast enhancement patterns that may indicate plaque inflammation. Measurement of carotid plaque signal intensity is limited by the non-linear relationship between MR signal intensity and the concentration and dynamics of gadolinium-based contrast agent distribution into the extracellular space. (5) Hence, normalizing plaque signal intensity to a concurrently acquired reference tissue, such as adjacent skeletal muscle, may provide a physiologically grounded approach to reduce inter-scan and inter-patient variability while preserving meaningful differences in contrast enhancement. (7, 8) DCE-MRI measures, including normalised signal intensity in the vessel wall and plaque core, may serve as surrogate markers for plaque physiological processes, offering insights into plaque composition, increased permeability, and enhancement related to neovascularisation. (9) Compared with pharmacokinetic parameters requiring complex modelling (e.g., transfer constant K^trans^ and plasma volume fraction v_p_) (5), this simplified normalisation strategy may prove more feasible, reproducible, and accessible, facilitating longitudinal assessment of contrast enhancement patterns potentially linked to plaque inflammation and vulnerability over time.

The primary aim of this exploratory study was to evaluate the reproducibility of normalised signal intensity (SI_norm_) measurements of plaque core and remote atherosclerosis-free carotid vessel wall as a potential surrogate marker of plaque neovascularisation. Secondary aims were to determine whether SI_norm_ in the plaque core differed from that in the remote carotid vessel wall. An additional secondary aim was to explore whether changes in normalized plaque core SI_norm_ from baseline to 6-month follow-up differed between participants receiving colchicine therapy and those receiving placebo.

## METHODS

### Study design

This retrospective longitudinal cohort study evaluated the feasibility and reproducibility of DCE-MRI plaque signal intensity measurements normalised to skeletal muscle as a surrogate marker of carotid plaque neovascularisation.

### The CAPRI study

Participants were retrospectively selected from the CAPRI study, a single-center, double-blind, randomized controlled trial that enrolled patients with mild-to-moderate carotid atherosclerosis confirmed by Doppler ultrasound. All participants underwent carotid MRI, including DCE-MRI, at baseline and at 6-month follow-up. The primary aim of the CAPRI study was to evaluate the effects of low-dose colchicine therapy versus placebo over 6 months on carotid plaque volume measured using high-resolution morphological MRI sequences and circulating systemic inflammatory markers. The MRI morphological analysis of the carotid plaque had a neutral outcome, with no effect of colchicine therapy on carotid plaque volume after 6 months in participants receiving colchicine therapy. Given the availability of paired DCE-MRI datasets in addition to the high-resolution morphological sequences, the present exploratory sub-analysis aimed to assess a simplified, normalized signal intensity-based quantification method for DCE-MRI.

### Study setting

Participant enrolment and data collection were conducted at the Hospital Clínico Pontificia Universidad Católica de Chile, Santiago, Chile.

### Human research ethics approval

Institutional review board approval was granted to conduct the CAPRI study. Following the conclusion of the trial, an amendment of the existing human research ethics approval was granted to permit a retrospective analysis of images obtained during the trial (REF: 170511002).

A modification to the existing ethics approval for the CAPRI study was granted as ethics exemption from The University of Sydney to include a post-hoc exploratory analysis of DCE-MRI images obtained during the primary study.

### Inclusion/exclusion criteria

CAPRI recruited asymptomatic patients with mild-to-moderate carotid stenoses as per North American Symptomatic Carotid Endarterectomy Trial (NASCET) criteria confirmed at recruitment by Doppler ultrasound, who were already receiving guideline-directed antiplatelet and statin therapy and had lipid levels optimised in line with accepted guidelines for patients at high risk of atherosclerotic cardiovascular disease (low-density lipoprotein cholesterol <2.5mmol/L). (10)

In CAPRI, patients were excluded if they had a known hypersensitivity to colchicine therapy, significant renal impairment, or hepatic dysfunction. Patients with a history of major adverse cardiovascular or cerebrovascular events over the preceding 6-months prior to recruitment or who were due to undergo elective carotid endarterectomy during the duration of the study were excluded. Patients who had existing inflammatory conditions which would increase systemic inflammatory markers or were already taking colchicine therapy or other anti-inflammatory medications were also excluded. Finally, where patients were unable to undergo contrast-enhanced carotid MRI due to implanted ferromagnetic medical devices, claustrophobia or allergy to gadolinium-based contrast agents, they were also excluded from the study. After agreeing to participation in the study, patients were randomly assigned to receive low-dose colchicine therapy or placebo in a 1:1 fashion in permutated blocks of four, to ensure there were no differences in age between the two groups. Patients, investigators and trial administration staff were blinded to allocation throughout the trial. Investigators were only unblinded during the statistical analysis phase of the study. Venous blood samples were collected at recruitment to establish baseline inflammatory marker levels, including Interleukin-6 (IL-6), Interleukin-18 (IL-18) and vascular cell adhesion molecule-1 (VCAM-1).

Within the present study, patients from the CAPRI cohort were included if they completed both baseline and 6-month follow-up DCE-MRI imaging. Patients were excluded where it was determined by readers that their scans were affected by severe motion artefact or otherwise had an image quality that was inadequate for reliable DCE-MRI analysis. Lesions were not considered in the analysis if there was evidence of interval carotid endarterectomy or carotid stenting at 6-months compared to baseline, which may have been overlooked during recruitment. After reviewing all images in the dataset, the decision to exclude cases was made by consensus between readers [TR, NG].

### Image acquisition

Participants underwent high-resolution multi-contrast carotid DCE-MRI at baseline and 6-months using a 3T MRI scanner (Ingenia Gyroscan, Philips Healthcare, Eindhoven, Netherlands). A localiser scan was first performed to identify carotid landmarks which were used to ensure that the field of view remained consistent between the baseline and follow-up scans. Following three-dimensional (3D) turbo-spin echo (TSE) imaging, patients were administered 20mL of a gadolinium-based contrast agent with a concentration of 0.1mmol/mL, intravenously by hand injection (Gadavist^®^ [gadobutrol], Bayer AG. Leverkusen, Germany). Following contrast administration, patients underwent T1-weighted DCE-MRI. A total of 21 DCE-MRI images of selected carotid slices to include the whole plaque were obtained over 256 seconds at 12-second intervals. Table 1 shows the details of the MRI protocols used in CAPRI.

**Table 1:**
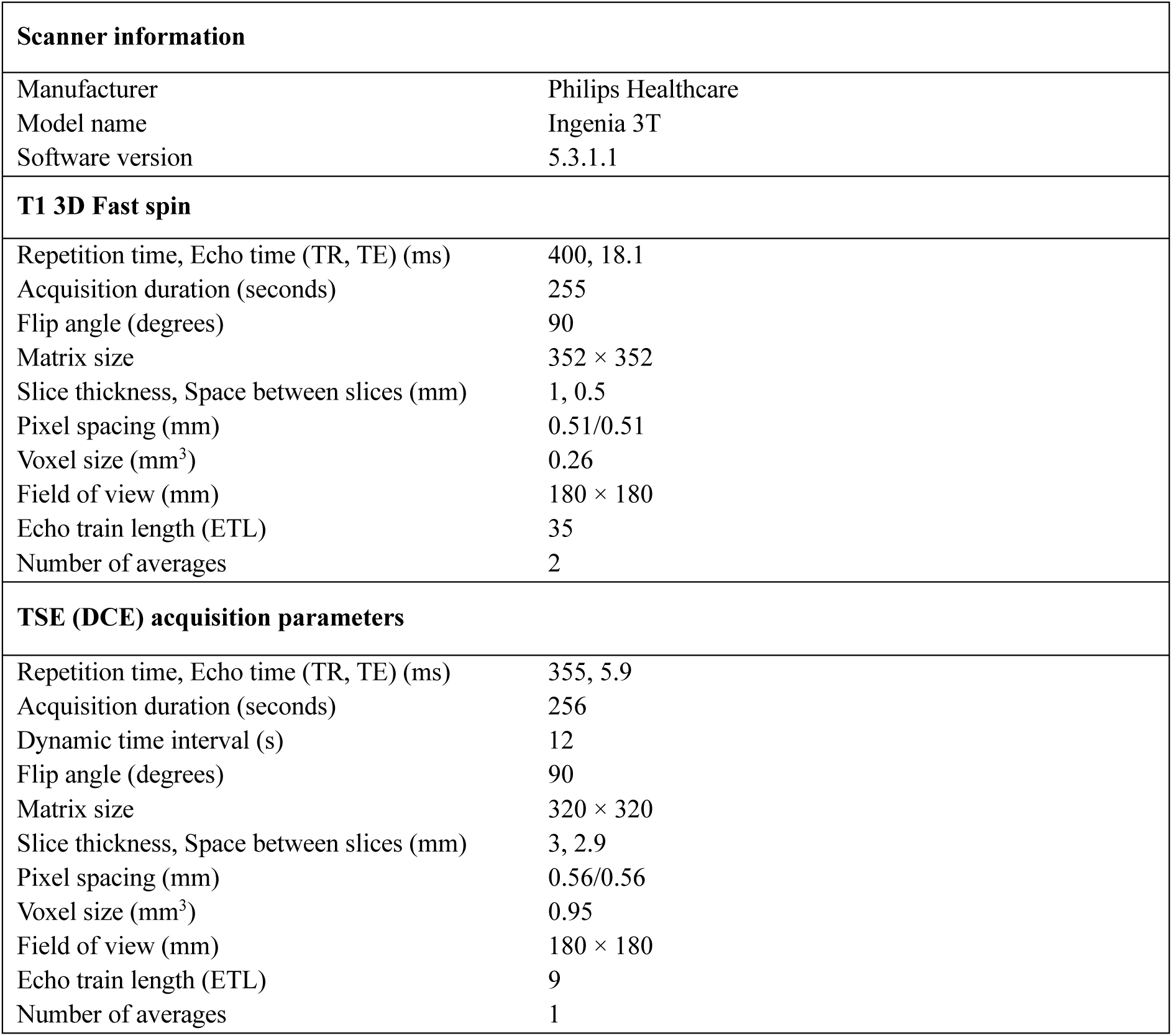
MRI parameters used for data acquisition.

| **Scanner information** |  |
| --- | --- |
| Manufacturer | Philips Healthcare |
| Model name | Ingenia 3T |
| Software version | 5.3.1.1 |
| **T1 3D Fast spin** |  |
| Repetition time, Echo time (TR, TE) (ms) | 400, 18.1 |
| Acquisition duration (seconds) | 255 |
| Flip angle (degrees) | 90 |
| Matrix size | 352 × 352 |
| Slice thickness, Space between slices (mm) | 1, 0.5 |
| Pixel spacing (mm) | 0.51/0.51 |
| Voxel size (mm<sup>3</sup>) | 0.26 |
| Field of view (mm) | 180 × 180 |
| Echo train length (ETL) | 35 |
| Number of averages | 2 |
| **TSE (DCE) acquisition parameters** |  |
| Repetition time, Echo time (TR, TE) (ms) | 355, 5.9 |
| Acquisition duration (seconds) | 256 |
| Dynamic time interval (s) | 12 |
| Flip angle (degrees) | 90 |
| Matrix size | 320 × 320 |
| Slice thickness, Space between slices (mm) | 3, 2.9 |
| Pixel spacing (mm) | 0.56/0.56 |
| Voxel size (mm<sup>3</sup>) | 0.95 |
| Field of view (mm) | 180 × 180 |
| Echo train length (ETL) | 9 |
| Number of averages | 1 |

### Image analysis

Images were analysed in Osirix MD V14.1.2 (Pixmeo, Geneva, Switzerland). Image analysis was conducted by a reader who was trained in carotid MRI segmentation and atherosclerotic plaque analysis and was blinded to patient allocation to colchicine therapy or placebo groups while completing the analysis [TR]. A sample (∼25%) of analysed cases were reviewed by two additional readers [NG, MU], who were also blinded to allocation, to confirm methodological approach and segmentation accuracy. Multi-planar reconstructions of the post contrast 3D T1-weighted sequences were used to identify the most-diseased slice in the atherosclerotic carotid artery, using the carotid bifurcation as an anatomical reference between baseline and follow-up. This facilitated accurate region-of-interest (ROI) placement. On the axial 3D T1 post contrast images at the level of the most-diseased slice, polygonal ROIs were manually delineated in three distinct areas as shown in figure 1: (1) the plaque core, (2) remote vessel, and (3) adjacent posterior neck skeletal muscle bundles (serving as the normalisation reference). **(Figure 1)**

**Figure 1:**
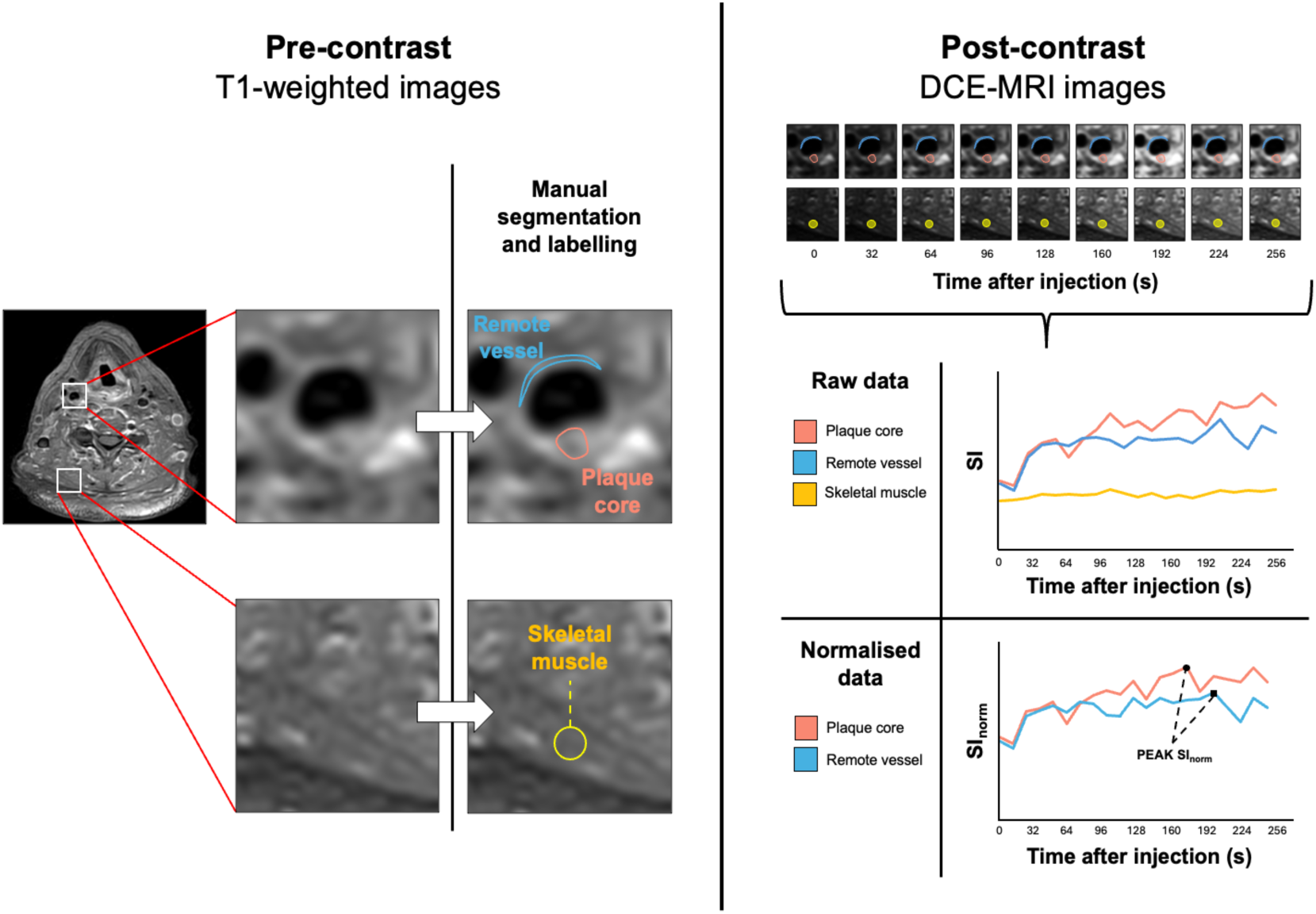
Segmentation method for evaluating signal intensity of carotid plaque core (pink), remote vessel (blue) and skeletal muscle (yellow). Top right show selected time frames from the 21 time frames of dynamic contrast-enhanced MRI acquisition.

Care was taken to ensure ROIs excluded the vessel lumen and adjacent tissues. The delineated ROIs from the 3D post-contrast T1-weighted images were then copied to the corresponding DCE-MRI series in the corresponding slice. Starting with the first frame of the DCE-MRI acquisition, ROIs were propagated semi-automatically to the subsequent 20 dynamic frames to collect the signal intensities from the three regions. Boundaries and positioning across the full DCE-MRI series were manually reviewed for each case and adjusted as needed on each frame to compensate for patient motion and carotid pulsation.

### Data processing

SI measurements from plaque core, remote carotid wall and skeletal muscle were exported from Osirix to MS Excel using a plugin tool (Export ROIs, V2.3). All SI measurements were first screened for outliers. Where the mean raw SI value of an ROI in one frame was more than three standard deviations from the mean of the same ROI in the preceding frame, the value was considered an outlier and was excluded from analysis. SI values from the plaque core and remote vessel wall ROIs were normalised independently at each dynamic time point by dividing the mean SI by the corresponding mean SI from a bundle of skeletal muscle in the posterior neck (splenius capitis). Parameters which were later used in the statistical analysis were derived from time-intensity curves of the normalised mean SI values (SI_norm_), using a custom-built Python script. The derived parameters were: (i) peak SI_norm_ (PEAK SI_norm_) and (ii) area-under-the-time-intensity-curve SI_norm_ (AUC SI_norm_). Absolute differences between baseline and 6-month SI_norm_ measurements were expressed as ΔSI_norm._

### Statistical analysis

Statistical analysis was performed using IBM SPSS Statistics (MacOS V29.0.2.0, SPSS Inc., Armonk, NY, USA). Data were determined as being normally or not normally distributed by using the Shapiro-Wilk test and visual inspection of the data distribution, and presented as mean ± SD or median [interquartile range] as appropriate. The paired T test, paired Wilcoxon test and Mann-Whitney U test were performed as appropriate. Spearman’s rho was calculated to assess correlations between continuous variables. A *p*-value <0.05 was considered statistically significant.

## RESULTS

A total of 28 carotid atherosclerotic lesions from 28 patients were analysed with DCE-MRI. **(Figure 2)**

**Figure 2:**
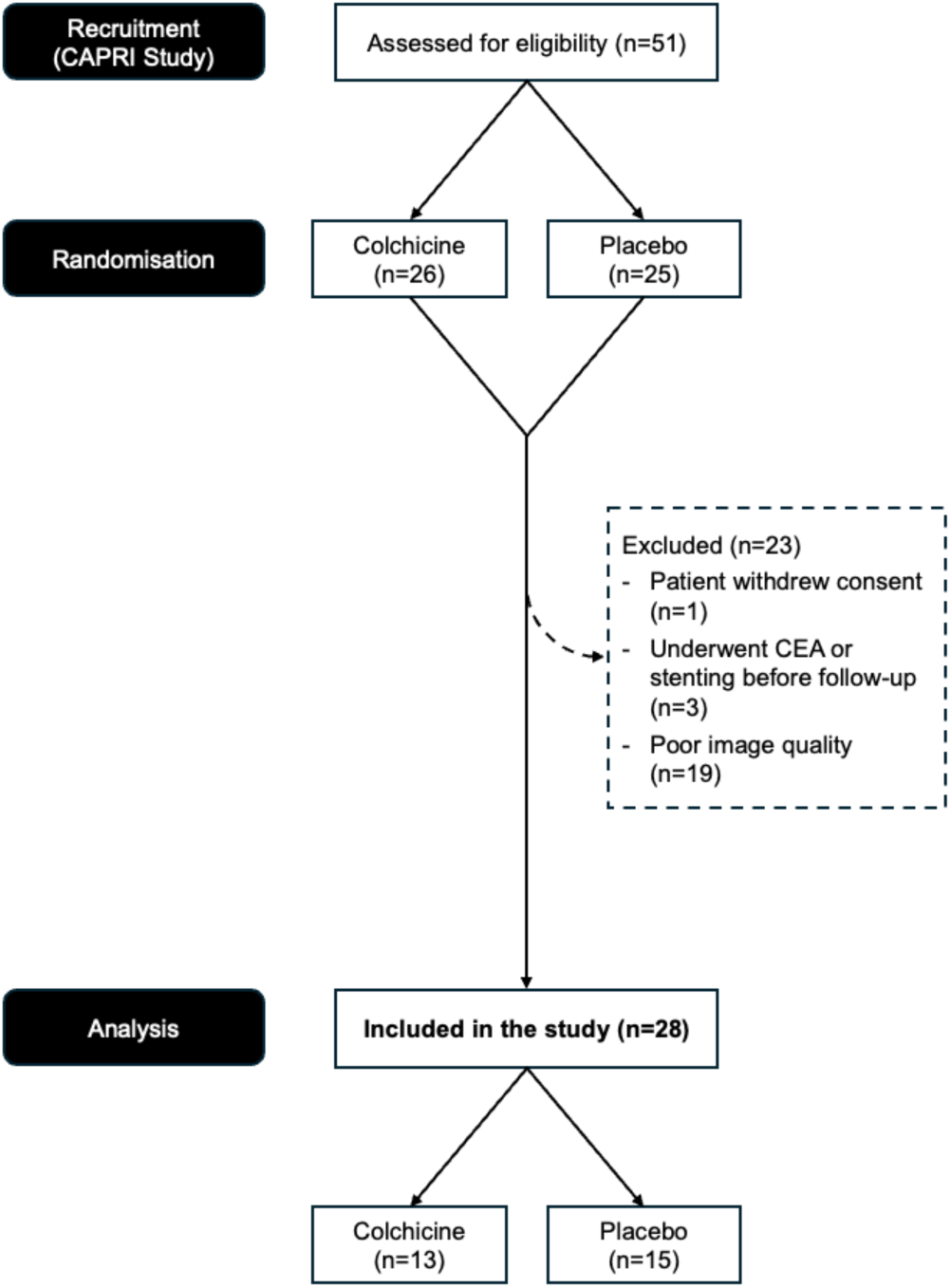
Flow chart of patient selection.

Of the 28 carotid lesions which were analysed in this study, 13 were from patients allocated to the treatment (colchicine therapy) arm (46%), and 15 were allocated to the placebo arm (54%). The median age of included patients was 72 [64-74] years. Of the 28 included patients, 10 (36%) were female. **(Table 2)**

**Table 2:** Patient characteristics at baseline.

| | All patients | Colchicine | Placebo | $p^{\dagger}$ |
| --- | --- | --- | --- | --- |
| **Number of subjects, n (row%)** | **28 (100)** | 13 (46) | 15 (54) |  |
| **Demographics** |  |  |  |  |
| Male, n (column%) | **18 (62)** | 8 (62) | 10 (67) | 0.78 |
| Female, n (%) | **10 (36)** | 5 (38) | 5 (33) |  |
| Age at enrolment, years | **72 [64-74]** | 64 [59-74] | 72 [66-74] | 0.51 |

| Clinical characteristics |  |  |  |  |
| --- | --- | --- | --- | --- |
| BMI, kg/m<sup>2</sup> | **27 [24-29]** | 26 [24-27] | 27 [25-29] | 0.41 |
| Diabetes, n (%) | **10 (36)** | 3 (23) | 7 (47) | 0.19 |
| Smoking, n (%) | **21 (75)** | 10 (77) | 11 (73) | 0.83 |
| Dyslipidaemia, n (%) | **26 (93)** | 11 (85) | 15 (100) | 0.12 |
| Hypertension, n (%) | **22 (79)** | 11 (85) | 11 (73) | 0.47 |
| Framingham 10-year MI risk, % | **19 [11-23]** | 15 [4-25] | 20 [17-22] | 0.46 |
| Moderate-to-Severe CAD, n (%) | **7 (25)** | 3 (23) | 4 (27) | 0.83 |
| Prior coronary revascularisation<sup>‡</sup>, n (%) | **14 (54)** | 9 (69) | 5 (33) | 0.26 |
| Prior MI, n (%) | **10 (38)** | 8 (62) | 2 (13) | **0.008** |
| Known PAD, n (%) | **6 (21)** | 2 (15) | 4 (27) | 0.47 |
| Prior cerebrovascular accident, n (%) | **2 (7)** | 1 (8) | 1 (7) | 0.92 |
| Family history of CAD, n (%) | **4 (14)** | 2 (15) | 2 (13) | 0.22 |
| Prescribed medications |  |  |  |  |
| SAPT/DAPT, n (%) | **26 (93)** | 13 (100) | 15 (100) | 0.62 |
| Beta blockers, n (%) | **14 (50)** | 7 (54) | 7 (47) | 0.71 |
| ACE inhibitors, n (%) | **22 (79)** | 11 (85) | 11 (73) | 0.47 |
| Statins, n (%) | **28 (100)** | 13 (100) | 15 (100) | 1.00 |
| Insulin or glucose-lowering agents, n (%) | **13 (46)** | 5 (38) | 8 (53) | 0.43 |
| All results expressed as *n* (column%) or Median [IQR] unless otherwise specified. |  |  |  |  |
| <sup>†</sup>Chi-square test, Significance level *p* <0.05 |  |  |  |  |
| <sup>‡</sup>Prior revascularisation includes prior drug-eluting stents or coronary artery bypass grafting |  |  |  |  |
| CAD –coronary artery disease |  |  |  |  |
| PAD – peripheral artery disease |  |  |  |  |
| MI – myocardial infarction |  |  |  |  |
| SAPT/DAPT – single antiplatelet therapy/dual antiplatelet therapy |  |  |  |  |

Plaque core PEAK SI_norm_ and AUC SI_norm_ measurements were consistently greater than remote vessel wall SI_norm_ measurements at baseline and at 6-month follow-up (p<0.001 for all). There was no difference between PEAK SI_norm_ or AUC SI_norm_ measurements for the plaque core or remote vessel wall at 6-months compared to baseline (p>0.05 for all). **(Figure 3)**

**Figure 3:**
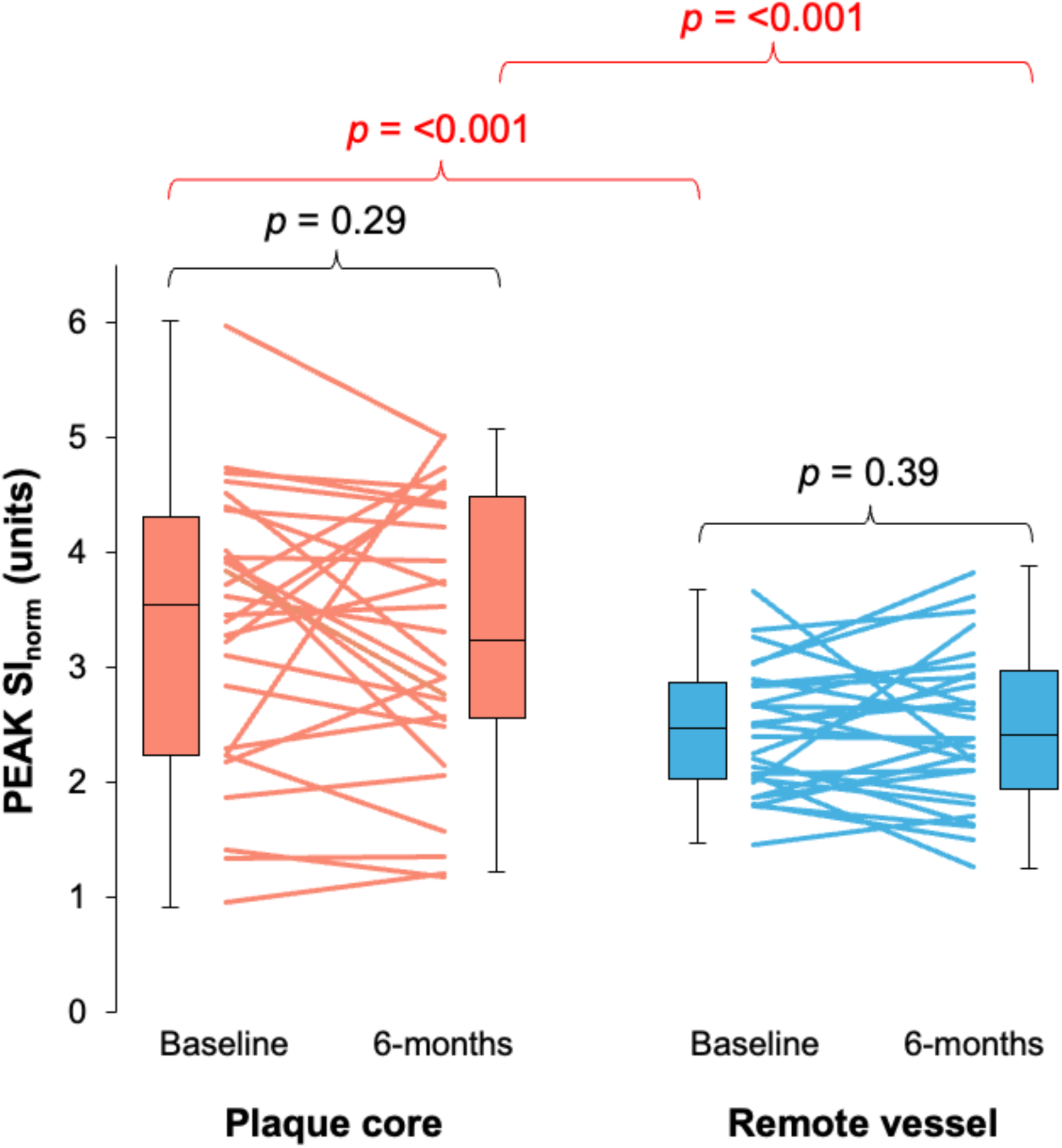
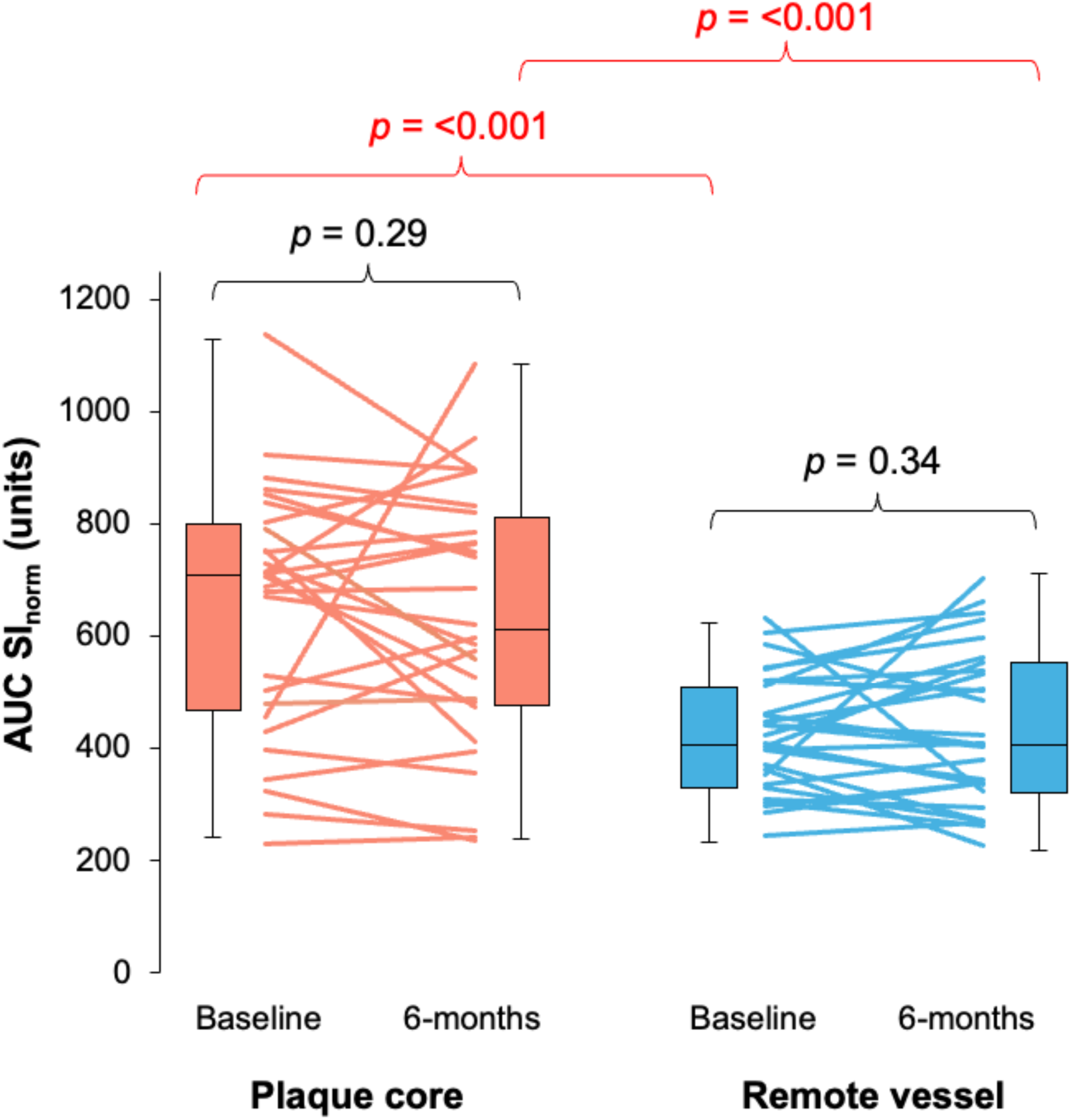
Paired line graph demonstrating changes in PEAK SI_norm_ (top) and AUC SI_norm_ (bottom) measurements in plaque core and remote vessel wall at baseline and 6-months.

The mean variance (ΔSI_norm_) for peak plaque core intensity at 6-months compared to baseline was −0.1 ± 0.96 units, while the mean variance for peak remote vessel wall intensity was 0 ± 0.55, which was not statistically significant, for either measure.

There were no correlations between plaque core peak SI_norm_ and circulating inflammatory biomarker levels, including hs-CRP, VCAM-1, IL-6 or IL-18 (*p*>0.05 for all). **(Figure 4)**.

**Figure 4:**
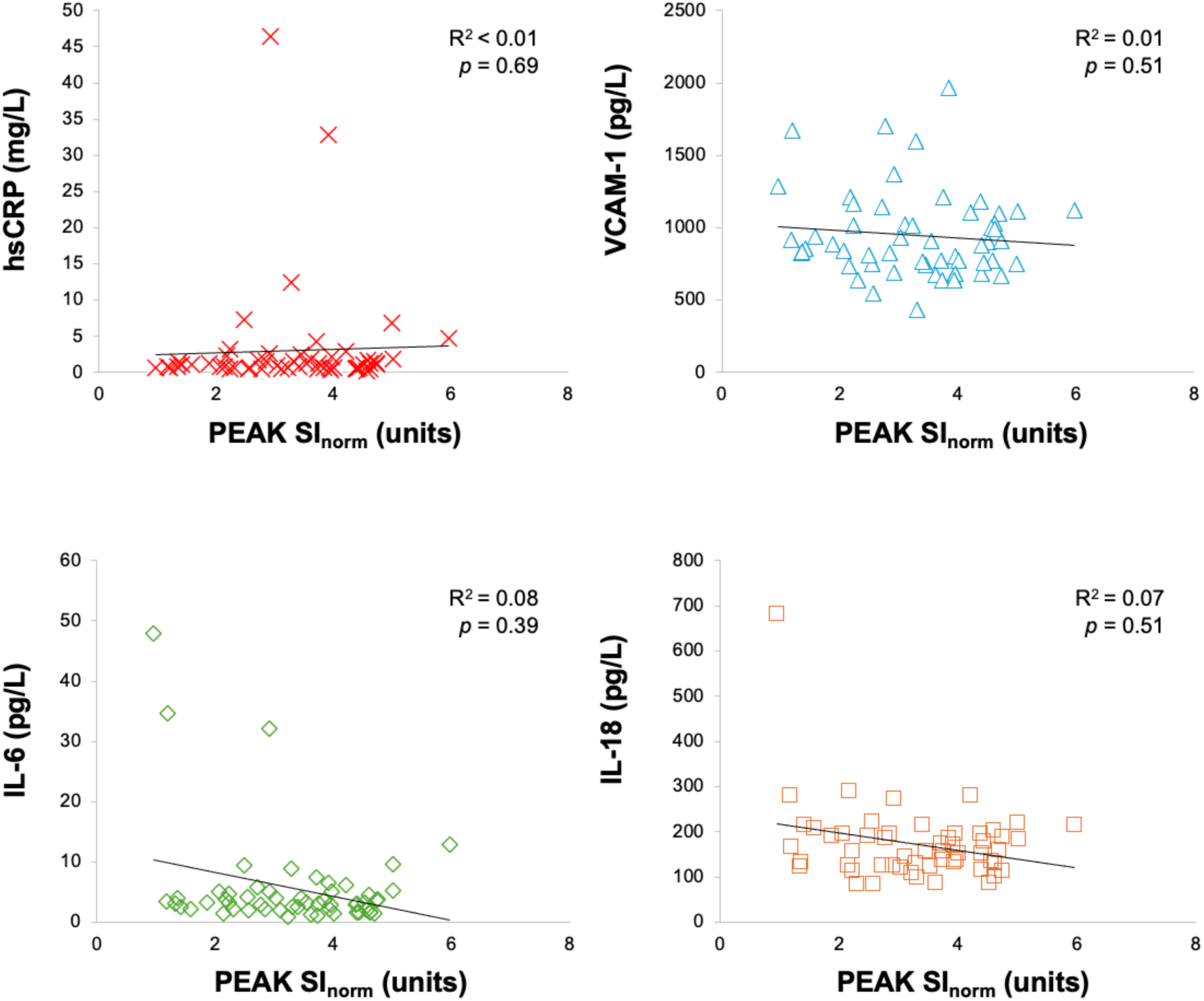
Correlations: Plaque core PEAK SI_norm_ and systemic inflammatory biomarker levels (clockwise from top left) hsCRP, VCAM-1, IL-18 and IL-6.

Finally, there were no differences in any changes in derived SI_norm_ parameters at baseline compared to 6-months in patients who received colchicine therapy and patients who received placebo, with median net changes in plaque core PEAK SI_norm_ of −0.3 [-1.0-0.2] versus −0.1 [-0.3-0.4] (Z = −0.94, *p* = 0.36, Mann-Whitney U test), plaque core AUC SI_norm_ of −49.8 [-142.5-13.3] versus 8.2 [-47.7-66.3] (Z = −1.1, *p* = 0.3), remote vessel wall PEAK SI_norm_ of 0.2 [-0.2-0.4] versus 0.0 [-0.5-0.3] (Z = −0.7, *p* = 0.48), and remote vessel wall AUC SI_norm_ of 35.8 [-39.8-54.6] versus 16.5 [-68.8-71.4] (Z = −0.9, *p* = 0.37), respectively. **(Figure 5)**

**Figure 5:**
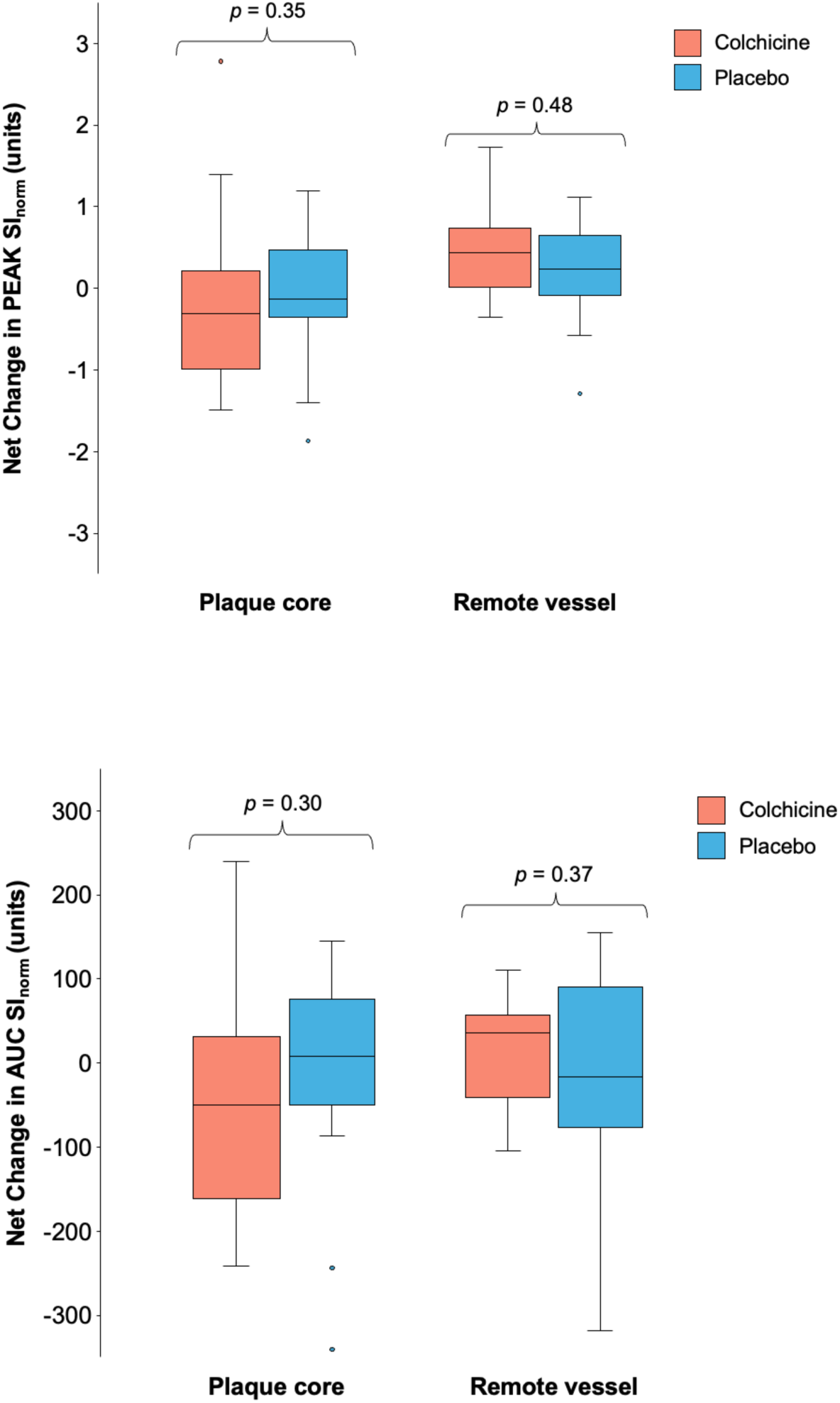
Net changes in PEAK SI_norm_ (top) and AUC SI_norm_ (bottom) between baseline and 6-month follow-up, colchicine vs. placebo groups.

## DISCUSSION

This exploratory DCE-MRI study showed that SI measurements of the carotid plaque core normalised to skeletal muscle are both reproducible in the same patient at 6-months, and differ from normalised measurements of remote vessel wall SI in regions of the carotid artery that were not affected by atherosclerosis. Pending future validation and head-to-head comparison with corresponding histological evidence and more advanced quantification parameters such as K^trans^, SI_norm_ demonstrated to be reproducible within the same patient over 6-months compared to baseline.

Metrics such as K^trans^ and v_p_ have been extensively used and physiologically validated as markers of plaque neovascularisation and microvascular permeability. However, they are limited by a lack of agreement regarding which pharmacokinetic model is most appropriate for atherosclerotic plaque evaluation, sensitivity to analysis parameters, and sensitivity to the choice of gadolinium-based contrast agent. (5, 9, 11) Additionally, some kinetic models of gadolinium contrast agents require arterial input functions, which are not able to be derived from black blood imaging. (5) To this end, SI_norm_ provides a more practical and accessible measure of DCE-MRI-derived enhancement, which is able to be drawn from black blood imaging without the need for an arterial input function. Its reproducibility also potentially makes SI_norm_ well-suited as a preliminary imaging biomarker for longitudinal studies where interscan comparability remains a challenge. Within the limitations of the present study, we found that there were no changes due to colchicine therapy on SI_norm_, and that there were no differences in change in SI_norm_ between colchicine and placebo groups.

### SI_norm_ as a surrogate marker of carotid plaque neovascularisation

SI_norm_ may represent a physiologically plausible semi-quantitative surrogate marker of carotid plaque neovascularisation. Neovessel-rich plaques are expected to show greater and more rapid enhancement due to increased microvessel density and capillary surface area. Hence, higher SI_norm_ values could reasonably be interpreted as reflecting increased plaque microvasculature. (6) The marked difference in SI_norm_ between the plaque core and the remote vessel wall observed in this study suggests that SI_norm_ has at least some sensitivity to atherosclerosis-specific physiological processes.

However, the biological specificity of SI_norm_ for carotid plaque neovascularisation remains incompletely characterised. It is possible that the metric incorporates multiple overlapping processes including changes in plaque perfusion, vascular permeability, and extracellular volume fraction, as opposed to neovascularisation alone. (12) Consequently, whilst SI_norm_ offers practical advantages in reproducibility and technical implementation for longitudinal studies, its mechanistic interpretation as a specific marker of neovascularisation should be done with caution, pending histopathological validation in larger, prospectively designed experiments.

It should also be noted that, because the segmentation technique relies on precise, manual discrimination between signal intensity in the plaque core and the surrounding carotid wall, a substantial number of cases (19/51, 37%) were excluded from the analysis due to patient motion or poor image quality, which prevented reliable identification of key vascular landmarks. This represents a practical limitation of the proposed method and may restrict its translatability to larger or more heterogeneous datasets.

### Temporal imaging in atherosclerosis evaluation

This study integrated a temporal dimension into the evaluation of carotid atherosclerosis by assessing both contrast enhancement-derived normalised signal intensity values immediately following gadolinium administration and longitudinal changes in normalised signal intensity over 6 months. Atherosclerosis is driven by multifaceted, dynamic physiological processes which includes inflammation, neovascularization, and altered perfusion, that may unfold concurrently or independently and that may be associated with plaque vulnerability and response to therapies. (13) Incorporating time-resolved imaging, as demonstrated here with DCE-MRI, moves beyond static morphological characterisation to capture evolving surrogate markers of contrast agent perfusion within plaques. Emerging imaging techniques such as dynamic total-body positron emission tomography (TB-PET) may extend this temporal approach systemically, enabling whole-body, time-resolved visualisation of radiotracer uptake in atherosclerotic lesions. (14) Much like DCE-MRI that reveals contrast wash-in and wash-out potentially reflective of surrogate markers of microvascular dynamics in individual plaques, dynamic TB-PET may provide enhanced kinetic modelling of molecular targets (e.g., inflammation, plaque metabolism and macrophage activity) across the entire vascular tree, offering complementary insights into plaque progression, destabilization, and systemic atherosclerotic burden. (14, 15) Therefore, future investigations in atherosclerosis should consider the inclusion of dynamic imaging modalities to interrogate surrogate markers of pathophysiological processes associated with contrast perfusion and their potential modulation by anti-inflammatory or plaque-stabilising therapies.

### Potential future applications of Artificial Intelligence and SI_norm_

The rapid development of Artificial Intelligence (AI) models capable of advanced, fully automated vascular segmentation represents an exciting and rapidly evolving frontier in atherosclerosis imaging. (16, 17) However, automated segmentation and extraction of carotid plaque neovascularisation metrics such as K^trans^ and v_p_, to our knowledge, have not yet been reported. A plausible explanation for this gap is that these parameters require robust estimation of an arterial input function in addition to accurate vessel and plaque segmentation, substantially increasing the algorithmic and computational complexity of AI- based workflows. Within this context, SI_norm_, once experimentally validated, may represent a more pragmatic and scalable target for early AI applications, as it provides a surrogate measure of plaque neovascularisation without necessitating explicit arterial input function modelling.

### Limitations

The most significant limitation of this study is the use of a non-validated, albeit physiologically plausible, metric of contrast arrival within the carotid wall and plaque due to the lack of T1 mapping imaging which was not available in this analysis. Additionally, although the image analysis methodology was reviewed by multiple experienced readers, inter-reader reliability was not evaluated within the present study. The limited sample size and relatively high number of patients who were excluded could also be considered limitations. The study’s strengths lie in its rigorous methodology, which has been proven to be reproducible at two different time points within the same patients. Future studies could address the limitations of the present study by using a more homogenised cohort, and by including comparison with validated histological or quantitative measures of plaque neovascularisation.

## CONCLUSION

DCE-MRI normalised PEAK and AUC signal intensity was greater for plaque core compared to remote vessel wall, and both measurements are reproducible over six months. This simplified approach may facilitate future assessment of neovascularisation and plaque phenotypes in clinical practice.

## LIST OF ABBREVIATIONS

AI: artificial intelligence
AUC: area under the curve
DCE-MRI: dynamic contrast-enhanced magnetic resonance imaging
hsCRP: high-sensitivity c-reactive protein
IL-6: interleukin-6
IL-18: interleukin-18
k-trans: volume transfer constant
MACCE: major adverse cardiovascular and cerebrovascular events
NASCET: North American Symptomatic Carotid Endarterectomy Trial
SI: signal intensity
SI_norm_: normalised signal intensity
TB-PET: total-body positron emission tomography
v_p_: plasma volume fraction
VCAM-1: vascular cell adhesion molecule-1

## Human research ethics compliance statement

This study was a sub-study of the CAPRI clinical trial, conducted at the Universidad Católica Clinical Hospital in Santiago, Chile. Institutional review board approval was granted to conduct the CAPRI study. Following the conclusion of the trial, an amendment of the existing human research ethics approval was granted to permit a retrospective analysis of images obtained during the trial.

## Data availability statement

The authors will respond to reasonable requests for data sharing of deidentified clinical data and quantitative image analysis data which support the findings of the present study.

## Ethics

This study was approved by the local human subject research ethics committee and all subjects provided written informed consent.

## REFERENCES

1. Zheng KH, Schoormans J, Stiekema LC, Calcagno C, Cicha I, Alexiou C, et al. Plaque permeability assessed with DCE-MRI associates with USPIO uptake in patients with peripheral artery disease. JACC: Cardiovascular Imaging. 2019;12(10):2081–3.

2. Yuan J, Makris G, Patterson A, Usman A, Das T, Priest A, et al. Relationship between carotid plaque surface morphology and perfusion: a 3D DCE-MRI study. Magnetic Resonance Materials in Physics, Biology and Medicine. 2018;31(1):191–9.

3. Truijman MT, Kwee RM, van Hoof RH, Hermeling E, van Oostenbrugge RJ, Mess WH, et al. Combined 18F-FDG PET-CT and DCE-MRI to assess inflammation and microvascularization in atherosclerotic plaques. Stroke. 2013;44(12):3568–70.

4. Khalifa F, Soliman A, El-Baz A, Abou El-Ghar M, El-Diasty T, Gimel’farb G, et al. Models and methods for analyzing DCE-MRI: A review. Medical physics. 2014;41(12):124301.

5. Gaens ME, Backes WH, Rozel S, Lipperts M, Sanders SN, Jaspers K, et al. Dynamic contrast-enhanced MR imaging of carotid atherosclerotic plaque: model selection, reproducibility, and validation. Radiology. 2013;266(1):271–9.

6. Dong L, Kerwin WS, Chen H, Chu B, Underhill HR, Neradilek MB, et al. Carotid Artery Atherosclerosis: Effect of Intensive Lipid Therapy on the Vasa Vasorum—Evaluation by Using Dynamic Contrast-enhanced MR Imaging. Radiology. 2011;260(1):224–31.

7. Bane O, Hectors SJ, Wagner M, Arlinghaus LL, Aryal MP, Cao Y, et al. Accuracy, repeatability, and interplatform reproducibility of T1 quantification methods used for DCE-MRI: Results from a multicenter phantom study. Magnetic resonance in medicine. 2018;79(5):2564–75.

8. Ng CS, Wei W, Bankson JA, Ravoori MK, Han L, Brammer DW, et al. Dependence of DCE-MRI biomarker values on analysis algorithm. PloS one. 2015;10(7):e0130168.

9. Calcagno C, Mani V, Ramachandran S, Fayad ZA. Dynamic contrast enhanced (DCE) magnetic resonance imaging (MRI) of atherosclerotic plaque angiogenesis. Angiogenesis. 2010;13(2):87–99.

10. Lalor E. National Vascular Disease Prevention Alliance. Guidelines for the management of absolute cardiovascular disease risk. 2012. ISBN 978–0–9872830–1–6. https://strokefoundation.com.au/~/media…; 2012.

11. Kerwin WS, Zhao X, Yuan C, Hatsukami TS, Maravilla KR, Underhill HR, et al. Contrast-enhanced MRI of carotid atherosclerosis: dependence on contrast agent. Journal of Magnetic Resonance Imaging: An Official Journal of the International Society for Magnetic Resonance in Medicine. 2009;30(1):35–40.

12. Calcagno C, Lobatto ME, Dyvorne H, Robson PM, Millon A, Senders ML, et al. Three-dimensional dynamic contrast-enhanced MRI for the accurate, extensive quantification of microvascular permeability in atherosclerotic plaques. NMR Biomed. 2015;28(10):1304–14.

13. Libby P. Inflammation during the life cycle of the atherosclerotic plaque. Cardiovasc Res. 2021;117(13):2525–36.

14. Borja AJ, Rojulpote C, Hancin EC, Høilund-Carlsen PF, Alavi A. An update on the role of total-body PET imaging in the evaluation of atherosclerosis. PET clinics. 2020;15(4):477–85.

15. Calcagno C, Ramachandran S, Izquierdo-Garcia D, Mani V, Millon A, Rosenbaum D, et al. The complementary roles of dynamic contrast-enhanced MRI and 18F-fluorodeoxyglucose PET/CT for imaging of carotid atherosclerosis. European Journal of Nuclear Medicine and Molecular Imaging. 2013;40(12):1884–93.

16. Wang Y, Yao Y. Application of artificial intelligence methods in carotid artery segmentation: a review. IEEE Access. 2023;11:13846–58.

17. Chen L, Zhao H, Jiang H, Balu N, Geleri DB, Chu B, et al. Domain adaptive and fully automated carotid artery atherosclerotic lesion detection using an artificial intelligence approach (LATTE) on 3D MRI. Magnetic Resonance in Medicine. 2021;86(3):1662–73.

